# A Machine Learning Framework for Melting Curve Analysis: Sequential Binary Encoding and Dual-Model Error Mitigation

**DOI:** 10.1101/2025.11.09.25339864

**Authors:** Ping Tang, Jin Chen, Bo Zhao, Guanbin Zhang, Shuang Jian, Tao Deng, Dong Liang

**Author notes:** Corresponding authors. E-mail addresses (D. Liang). 1. Author Contributions: Conceptualization: Dong Liang, Ping Tang, Guanbin Zhang, Bo Zhao, Jin Chen, Tao Deng Data curation: Ping Tang, Dong Liang, Guanbin Zhang, Shuang Jian Methodology: Ping Tang, Dong Liang, Jin Chen, Bo Zhao, Guanbin Zhang Supervision: Ping Tang, Dong Liang, Tao Deng, Jin Chen, Bo Zhao Validation: Bo Zhao, Jin Chen, Ping Tang Visualization: Jin Chen, Bo Zhao, Ping Tang, Dong Liang Writing - original draft: Jin Chen, Ping Tang, Dong Liang, Shuang Jian Writing - review & editing: Jin Chen, Ping Tang, Dong Liang, Tao Deng.

## Abstract

As a cornerstone technique in molecular diagnostics, melting curve analysis (MCA) enables cost-effective multiplex detection using widely accessible fluorescent PCR instruments, eliminating the need for expensive sequence-specific probes. Nevertheless, the broader clinical application of MCA faces limitations due to several interpretation challenges, primarily concerning signal noise, baseline drift, and inter-operator variability.

To address these limitations, we developed a dual-model machine learning framework trained on 186,138 samples and validated with 25,918 independent samples. The first model performs curve quality control (QC) using a binary XGBoost classifier (500 trees, depth=10) to filter non-informative curves. The second model determines melting temperature (Tm) values via a 151-bit encoded vector spanning 40-85°C at 0.3°C resolution.

Internal validation demonstrated high accuracy of the framework in the automatic interpretation of MCA results directly from raw data. External validation showed strong concordance with manual interpretation, with 90.5% of discrepant cases supporting the framework’s predictions upon secondary expert review. Five-fold cross-validation on a balanced subset of 28,880 samples achieved an average accuracy of 98.45% (95% CI: 96.84%-100.00%) and an area-under-the-curve (AUC) value of 0.9991 (SD +/- 0.0012). The system maintained consistent performance across four fluorescence channels (FAM/VIC/ROX/CY5) and substantially reduced interpretation time compared with manual methods.

In summary, this work establishes a robust and scalable strategy for the automated interpretation of MCA. The proposed framework can be readily integrated with existing PCR platforms, paving the way for standardized, high-throughput, and intelligent MCA-based molecular diagnostics.

## 1. Introduction

Melting curve analysis (MCA) has become a fundamental technique for assessing amplicon specificity and detecting sequence polymorphisms, eliminating the need for sequence-specific probes. As the temperature gradually rises, double-stranded DNA (dsDNA) undergoes thermal denaturation, releasing intercalating fluorescent dyes such as SYBR Green or EvaGreen, which causes a corresponding decrease in fluorescence intensity [1, 2]. Plotting fluorescence against temperature generates the characteristic melting curve, and its first derivative (–dF/dT) reveals the melting temperature (Tm)—the point at which 50% of dsDNA molecules denature. Tm values, determined by sequence length, GC content, and complementarity, act as molecular fingerprints for distinguishing genotypes, detecting pathogens, or identifying genetic variants. High-resolution MCA (HR-MCA), also known as HRM, is made possible by precise temperature control and dense signal acquisition, achieving single-nucleotide polymorphisms (SNPs) resolution. This technique has been successfully applied in molecular subtyping, antimicrobial resistance gene detection, and viral genotyping [2].

MCA offers several key advantages, including low cost, no requirement for labeled probes, and high throughput, making it particularly suitable for multiplex detection in large-scale diagnostic workflows [3]. Nevertheless, traditional MCA interpretation continues to rely heavily on manual inspection or proprietary instrument software, both of which are susceptible to baseline drift, signal fluctuation, and operator bias. These limitations are further amplified in heterogeneous clinical samples, where overlapping peaks, multiple amplicons, or minor sequence variants complicate accurate Tm determination [4]. Furthermore, variations in thermal ramping protocols, optical detection systems, and inconsistencies in data formats across qPCR platforms create significant barriers to cross-laboratory reproducibility and data integration, hampering MCA’s standardization in clinical practice.

Recent advancements in machine learning (ML) have opened up new avenues for mitigating these limitations [5, 6], particularly through the automated extraction of complex curve features such as peak shape, slope changes, and Tm distribution patterns, which in turn enhance classification robustness and Tm prediction accuracy. Previous studies have shown that algorithms such as support vector machines, dynamic time warping, and neural networks can improve the sensitivity and reproducibility of MCA-based assays [7]. Nonetheless, critical gaps persist — large-scale clinical validation, multi-channel performance assessment, and systematic exploration of cross-platform generalization (notably including challenges posed by incompatible data standards)—have yet to be fully addressed.

In this study, we explored the potential of a dual-model framework based on the XGBoost algorithm [8] for automated melting curve interpretation in the molecular diagnosis of thalassemia. The first model employs a binary classifier to identify valid Tm peaks (‘no Tm’ vs. ‘has Tm’), while the second model classifies positive cases into specific Tm categories. We leveraged a large dataset comprising over 210,000 raw melting curves across multiple fluorescence detection channels. Our validation strategies included internal testing, independent external validation, and the use of balanced five-fold cross-validation on a representative subset. Additional analyses investigated channel-specific performance variability, the impact of training data size and class imbalance, and practical challenges encountered in cross-platform data integration, noting that inconsistencies in signal normalization, data formatting, and melting curve profiles across different qPCR platforms contribute to the observed difficulty of cross-platform generalization. This work aims to document these challenges and provide insights into the complexity of developing reproducible and scalable solutions for high-throughput, Tm-based molecular diagnostics.

**Scheme 1.**
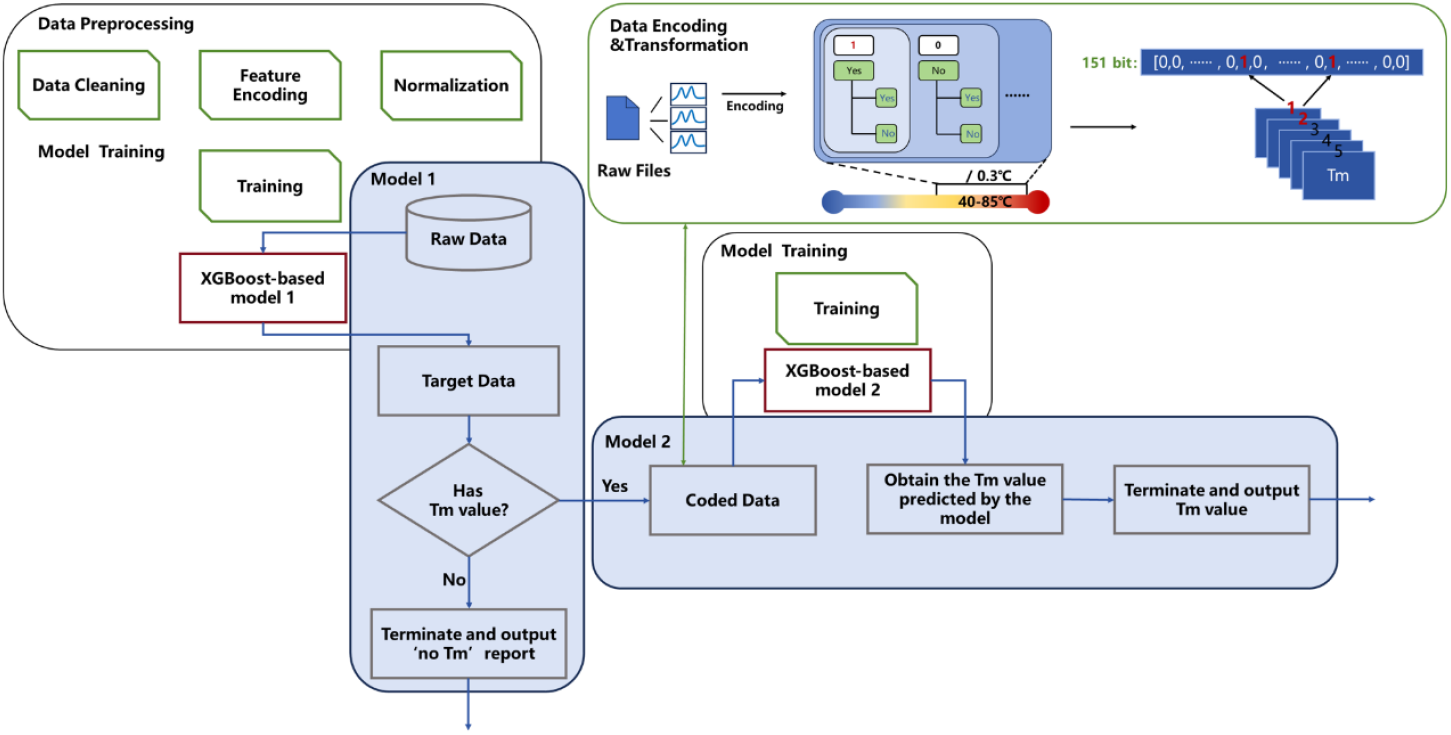
Conceptual workflow integrating data preprocessing, model training, and decision-making steps. The process starts with data preprocessing and proceeds to training ‘Model 1’. Raw data are converted into target data, which are subsequently checked for the presence of a Tm value. If no Tm is detected, the workflow ends by reporting ‘no Tm’. If a Tm is present, the resulting coded data serve as input to ‘Model 2’, which determines the specific Tm value—covering a temperature range of 40–85°C with a precision of ±0.3°C. The workflow concludes by outputting the predicted Tm value. In the coding step, the 40–85°C temperature range is partitioned into 151 intervals (each 0.3°C), and Tm values are converted into 151-bit binary vectors.

## 2. Methods

### 2.1 Melting Curve Morphology and Analytical Implications

Three characteristic melting profiles are typically observed in MCA: homogeneous, heterogeneous, and failure. A homogeneous profile, characterized by a single sharp peak, indicates successful amplification of the target region with wild-type alleles (i.e., a single PCR product). A heterogeneous profile, exhibiting multiple peaks, suggests the presence of mutant alleles in the target region (i.e., PCR products of multiple sequence variants), while minor non-specific amplification may appear as background noise. A failure profile, marked by the absence of peaks, signifies a failed amplification (i.e., no PCR product). These morphological features are critical for Tm-based applications [9, 10], including genotyping [11], multiplex pathogen detection, and troubleshooting.

### 2.2 Datasets

A total of 212,056 raw melting curve samples generated by a single fluorescent PCR instrument (SLAN-96S, Shanghai Hongshi Medical Technology Co., Ltd, Shanghai, China) were collected from thalassemia-related mutation assays. Among these, 186,138 samples were used for model training and internal validation, and 25,918 were reserved for external validation. The data covered multiple fluorescence detection channels, specifically FAM, VIC, ROX, and CY5. Each record included up to five Tm values, along with raw melting curve data. In addition, an independent dataset (4,158 samples included) generated by another qPCR platform (Gentier 96R, Xi’an Tianlong Science and Technology Co., Ltd, Xi’an, China) was collected for cross-platform validation. This dataset was used solely to evaluate cross-platform generalizability.

### 2.3 Data Preprocessing

For the first model, samples lacking valid Ct values (indicating failed experiments) were excluded. For the second model, building on the result of the first model, samples labeled as ‘no Tm’ were discarded. Each dataset retained only essential features: normalized melting curve data and up to five Tm values.

All data processing was performed using Python 3.12.

### 2.4 Tm Values Feature Encoding

To generate a structured representation of Tm peaks compatible with machine learning algorithms, a binary vector encoding scheme was employed. The temperature range of 40.0–85.0 °C was partitioned into 151 discrete intervals with a step size of 0.3 °C, matching the native output resolution of the SLAN-96S instrument. Finer temperature bins could be utilized if needed. Each interval is mapped to a specific temperature (e.g., bit 1 = 40.0 °C, bit 2 = 40.3 °C, …, bit 151 = 85.0 °C). For each extracted Tm value, the nearest temperature bin was assigned a value of ‘1’, with all other bins set to ‘0’, generating a structured binary vector suitable for machine learning input.

### 2.5 Model Architecture

The modeling framework comprised two sequential XGBoost classifiers. The first was a binary classifier designed to distinguish between curves with valid Tm peaks and those without, thereby filtering out failed or abnormal amplification reactions. The second was applied only to samples flagged as ‘has Tm’ by the first classifier. It consisted of 151 parallel binary classifiers, each predicting the presence or absence of a Tm peak within a specific temperature bin, thus framing the task as a multi-label classification problem. Both models configured with identical hyperparameters: 500 decision trees, a maximum tree depth of 10, and a learning rate of 0.05. Each classifier used binary logistic regression loss to model the probability of Tm peak presence, yielding independent probability outputs for all temperature bins.

### 2.6 Validation Strategy

Model validation employed a multi-layered strategy to assess predictive accuracy, robustness, and generalizability comprehensively.

We first designated 27,466 samples from the internal dataset as a hold-out test set, and model predictions were directly compared to the original PCR instrument outputs. Discordant cases between model predictions and instrument outputs underwent independent review to inform further analysis [12].

To characterize the natural variation of Tm peaks, we next analyzed Tm value distribution across the internal dataset. This analysis revealed a strong concentration of peaks around 67.3°C, consistent with the predominant assayed genotype target in thalassemia detection. The observed distribution not only characterized the underlying biological signal but also informed the design of subsequent validation experiments.

Based on this distribution, we constructed a balanced subset of 14,440 positive and 14,440 negative samples, centered on the dominant 67.3°C Tm peak. The subset was partitioned into five folds, each serving once as the test set. This five-fold cross-validation enabled systematic evaluation of model stability, with performance quantified via Accuracy, Precision, Recall, F1-score, AUC, and calibration metrics (RMSE, MAE), thereby assessing both decision-level accuracy and probability-level reliability.

To further examine generalizability, we performed external validation using an independent dataset of 25,918 samples obtained from separate experimental runs on the same qPCR platform. We paid particular attention to cases where the instrument indicated a Tm but the model did not, and these cases underwent manual reassessment to clarify whether discrepancies stemmed from borderline peaks or instrument miscalls.

In addition, we stratified the internal validation results by fluorescence detection channel (FAM, VIC, ROX, CY5) to determine whether optical differences influenced predictive performance. Melting curve data from each channel were independently extracted, with the model tested under identical data processing and validation conditions.

To probe the effect of training data size and class distribution on model performance, we conducted two independent experiments. In the first, balanced training sets of increasing sizes (200, 1,000, 2,000, 10,000, and 20,000 samples) were used to evaluate performance scalability. In the second, the number of positive samples was fixed at 10,000, while the number of negatives varied (100, 500, 1,000, 5,000, and 10,000), allowing assessment of class imbalance.

For benchmarking, we further compared XGBoost against seven alternative models: Random Forest, LightGBM, CatBoost, Logistic Regression, deep neural network (DNN), convolutional neural network (CNN), and recurrent neural network (RNN) [13, 14]. All models were trained and evaluated with identical feature sets, data splits, and a comparable hyperparameter tuning budget.

To evaluate cross-platform generalizability, we conducted validation using melting curve datasets generated from a second qPCR instrument (Gentier 96R). Specifically, XGBoost models trained exclusively on SLAN-96S data were directly applied to Gentier 96R datasets, without any retraining, recalibration, or normalization. Meanwhile, models trained on Gentier 96R data were respectively tested on Gentier 96R and SLAN-96S datasets. This dual validation strategy enabled assessment of both intra-platform and cross-platform performance, revealing how instrument-specific signal characteristics impact the robustness of Tm prediction models across different qPCR instruments.

## 3. Results

### 3.1 Internal Independent Validation

For internal validation, 27,466 samples were analyzed, taking the original PCR instrument’s Tm presence/absence calls as the reference standard. Comparisons of the model’s predictions against the instrument’s outputs showed the following (Figure 1a): For cases where the PCR instrument reported ‘no Tm’, the model predicted ‘no Tm’ for 940 cases and classified 87 cases as ‘has Tm’. For cases where the instrument reported ‘has Tm’, the model predicted ‘has Tm’ for 26,408 cases and classified 31 cases as ‘no Tm’.

**Figure 1.**
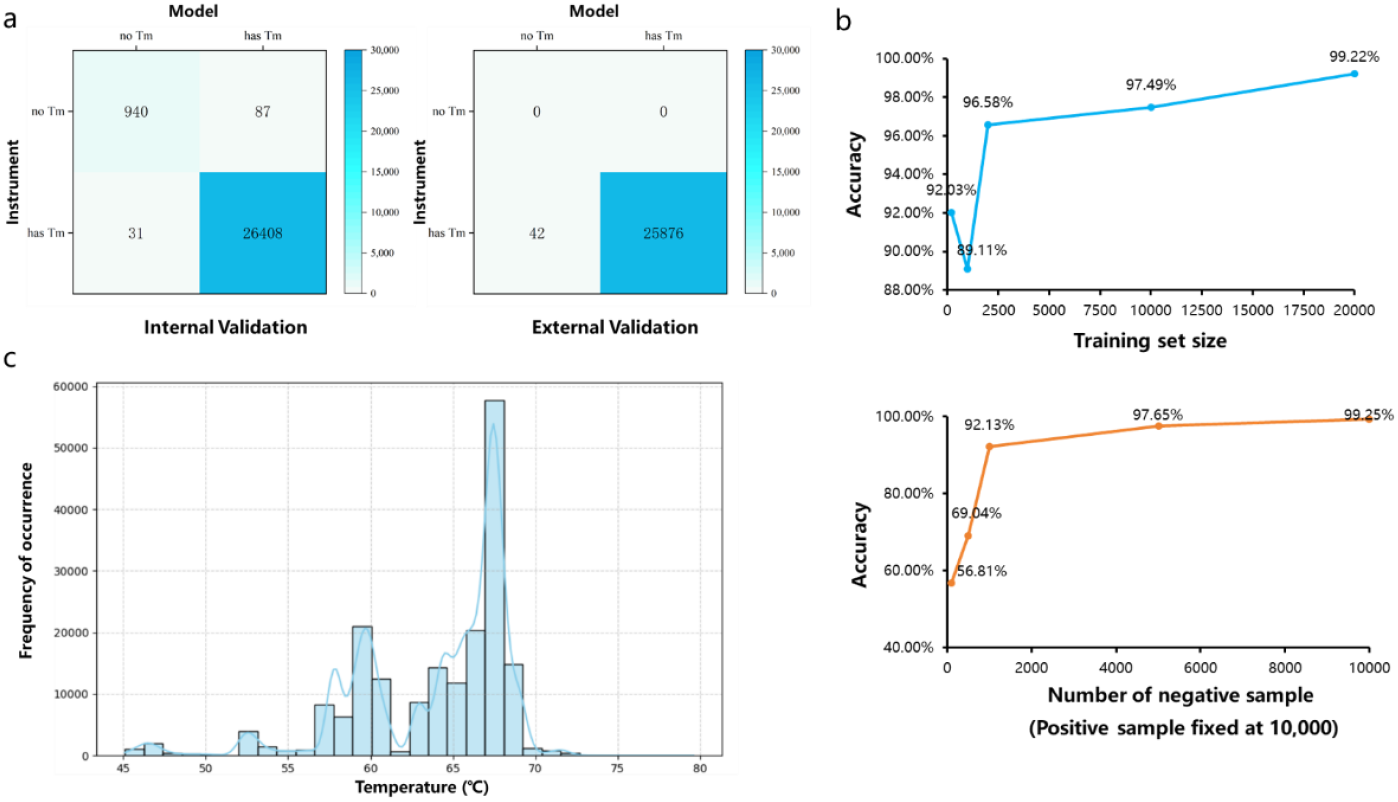
(a) Confusion matrix for internal validation and confusion matrix for external validation. (b) Effects of training size and class imbalance. Accuracy improved with larger balanced datasets and plateaued beyond 20,000 samples, while imbalance degraded performance. The accuracy drop at the second data point in the training set size experiment may stem from class imbalance (underrepresentation of negative samples) at this sample size, resulting in a reduction in the model’s performance. (c) Distribution of Tm values across datasets, with a dominant peak near 67.3°C corresponding to the target amplicon, and additional minor peaks that may reflect non-specific amplification, allelic variants, or experimental variability.

To further evaluate the model’s prediction accuracy, all discordant cases underwent independent review. Among the 87 cases where the instrument reported ‘no Tm’ but the model predicted ‘has Tm’, 55 were confirmed to contain valid Tm peaks, indicating that the model had correctly identified Tm presence, even though the instrument had returned a negative call. Conversely, among the 31 cases where the instrument reported ‘has Tm’ but the model predicted ‘no Tm’, 29 were confirmed to lack valid Tm peaks.

These findings demonstrate that the model not only aligned with the PCR instrument’s call in most cases but also corrected a significant number of apparent misclassifications, thereby reducing both false negatives and false positives in Tm detection.

### 3.2 Tm Distribution Analysis

Analysis of the internal dataset revealed that Tm peaks were not uniformly distributed but clustered around several dominant regions. The most prominent and clinically relevant cluster was centered near 67.3°C, a temperature that corresponds to the expected Tm peak of thalassemia-associated amplicons. This understanding of Tm distribution informed the design of subsequent cross-validation experiments, ensuring that validation subsets adequately represented the dominant Tm region (Figure 1c).

### 3.3 Five-Fold Cross-Validation on Internal Data

To assess the classifier’s stability under balanced conditions, specifically within the representative 67.3°C peak region, we used a subset of 14,440 positive and 14,440 negative samples for five-fold cross-validation. Across the five folds, the model achieved an average accuracy of 98.45% (95% CI: 96.84– 100.00%) and an AUC of 0.9991 (SD ± 0.0012). Fold-wise accuracy ranged from 96.10% to 99.27%. Additional performance metrics included Precision (98.63%), Recall (98.27%), and F1-score (98.43%), consistently reflecting high classification reliability. Calibration metrics also showed low error (RMSE: 0.1176 ± 0.0455; MAE: 0.0155 ± 0.0133), validating the stability of probability estimates across all folds (Table 1).

**Table 1.** Five-fold cross-validation performance on balanced internal subset.

| Fold | Accuracy | Precision | Recall | F1-score | AUC | RMSE | MAE |
| --- | --- | --- | --- | --- | --- | --- | --- |
| 1 | 0.961 | 0.9886 | 0.9328 | 0.9599 | 0.997 | 0.1974 | 0.039 |
| 2 | 0.9903 | 0.9849 | 0.9958 | 0.9904 | 0.9996 | 0.0985 | 0.0097 |
| 3 | 0.9927 | 0.9873 | 0.9983 | 0.9928 | 0.9997 | 0.0853 | 0.0073 |
| 4 | 0.9877 | 0.9865 | 0.9889 | 0.9877 | 0.9994 | 0.1109 | 0.0123 |
| 5 | 0.9908 | 0.9843 | 0.9976 | 0.9909 | 0.9996 | 0.0958 | 0.0092 |
| Avg | 0.9845 | 0.9863 | 0.9847 | 0.9843 | 0.9991 | 0.118 | 0.0155 |

### 3.4 External Independent Validation

The external validation set comprised 25,918 samples. We also compared the model’s predictions with outputs from the original PCR instrument. For cases labeled “no Tm” by the instrument, the model produced matching “no Tm” predictions. For cases where the instrument reported ‘has Tm,’ the model predicted ‘no Tm’ for 42 cases and ‘has Tm’ for 25,876 cases. Manual review of the 42 discordant cases showed that 38 were correctly classified as lacking valid Tm peaks, yielding an accuracy of 90.5% for this subset. These results demonstrate that the model maintained high concordance with the instrument’s outputs on external datasets and improved robustness in resolving ambiguous or borderline melting curves (Figure 1a).

### 3.5 Performance Across Fluorescence Channels

When stratified by fluorescence channel (CY5, FAM, ROX, VIC), performance differences were minimal. This variation was largely driven by training sample availability rather than intrinsic optical differences. All channels achieved high accuracy (>97.5%) and AUC (>0.994), which confirms that the model generalized effectively across different optical channels (Table 2).

**Table 2.** Channel-specific performance on internal validation set.

| Channel | Accuracy | Precision | Recall | F1-Score | AUC |
| --- | --- | --- | --- | --- | --- |
| CY5 | 0.9755 | 0.9067 | 0.9943 | 0.9485 | 0.9986 |
| FAM | 0.9918 | 0.9901 | 0.9982 | 0.9941 | 0.9997 |
| ROX | 0.9988 | 1.0000 | 0.8889 | 0.9412 | 0.9950 |
| VIC | 0.9932 | 0.9883 | 0.9980 | 0.9931 | 0.9999 |

### 3.6 Training Size and Class Imbalance Effects

From controlled experiments, we found that increasing the size of a balanced training set improved accuracy, and accuracy gains plateaued once the set exceeded 20,000 samples. Severe class imbalance, particularly when negative samples were underrepresented, led to a substantial decrease in accuracy. Restoring this balance recovered performance to near-optimal levels, emphasizing the importance of representative sampling for robust model application (Figure 1b).

### 3.7 Baseline Model Comparison

To benchmark the proposed performance, we compared the proposed XGBoost model against Random Forest, LightGBM, CatBoost, Logistic Regression, and deep learning models (DNN, CNN, RNN) under identical training and validation conditions. All models achieved high accuracy in Tm prediction under identical data splits (Table 3). However, XGBoost consistently demonstrated superior stability and robustness, making it well-suited for integration into the automated diagnostic workflow.

**Table 3.** Performance comparison of baseline models on Tm prediction.

| Model | Accuracy | Precision | Recall | F1-Score |
| --- | --- | --- | --- | --- |
| CatBoost | 0.9893 | 0.9813 | 0.9976 | 0.9894 |
| LightGBM | 0.9907 | 0.9843 | 0.9972 | 0.9907 |
| LogisticRegression | 0.9545 | 0.9187 | 0.9972 | 0.9563 |
| RandomForest | 0.9900 | 0.9829 | 0.9972 | 0.9900 |
| XGBoost | 0.9908 | 0.9843 | 0.9976 | 0.9909 |
| DNN | 0.9792 | 0.9641 | 0.9955 | 0.9796 |
| CNN | 0.9860 | 0.9743 | 0.9983 | 0.9861 |
| RNN | 0.9527 | 0.9206 | 0.9910 | 0.9545 |

### 3.8 Cross-Platform Validation

During preprocessing, we observed that although the peak positions of melting curves showed high consistency across platforms after expert calibration, the baseline regions before and after the peaks differed substantially (Figure 2). These discrepancies might contribute to reduced performance when applying models across instruments. Comparative visualization confirmed that Gentier 96R curves displayed elevated baseline noise and altered slopes relative to SLAN-96S data, despite the alignment of their peak temperatures around 67.3°C.

**Figure 2.**
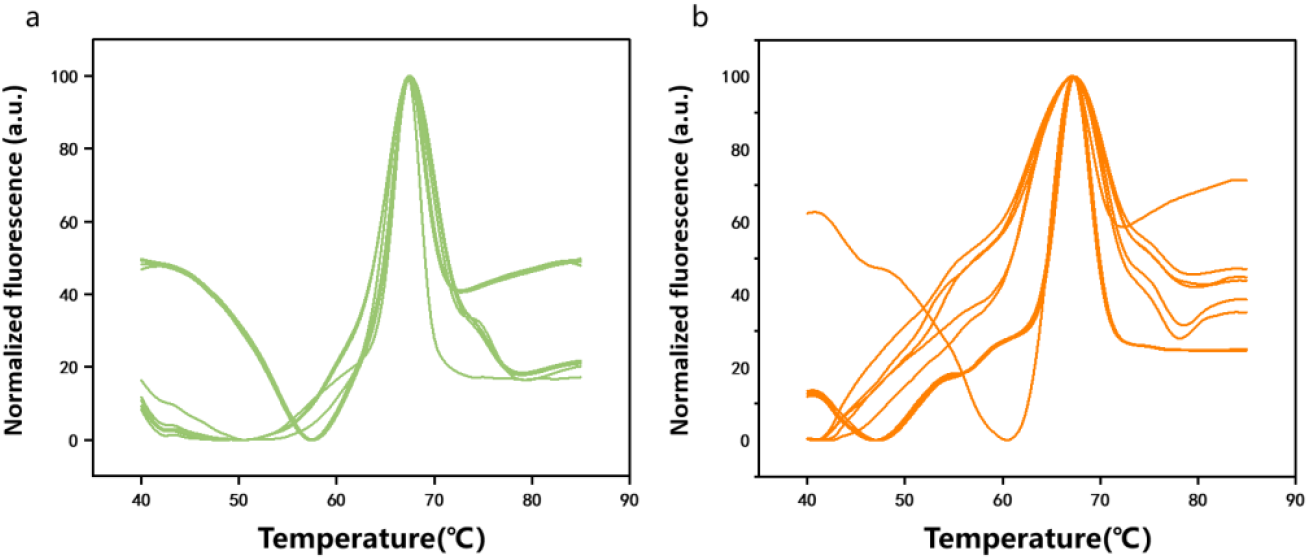
Representative melting curves from two qPCR platforms. From (a) SLAN-96S and (b) Gentier 96R instruments. Peaks align near 67.3°C, but baseline shapes differ, showing higher noise and altered slopes in Gentier 96R data.

When the model trained on SLAN-96S data was directly applied to Gentier 96R datasets, overall accuracy dropped to 20.34%, indicating poor generalizability without retraining or normalization. In contrast, the model trained and tested within the Gentier 96R platform achieved 74.30% accuracy, confirming that platform-consistent training preserved predictive reliability. Conversely, when the model trained on Gentier 96R data was applied to SLAN-96S test data, the accuracy was 34.53%, again reflecting asymmetric transferability between systems.

Manual inspection of discordant predictions showed that the majority of errors originated from differences in baseline characteristics rather than shifts in Tm peak temperatures. These results suggest that while the model framework performs robustly within individual platforms, successful cross-platform deployment requires baseline harmonization or multi-platform training to enhance generalizability across heterogeneous qPCR instruments.

## 4. Discussion

In this study, we present a dual-model XGBoost-based analytical framework that achieves high accuracy in automated Tm detection and prediction from melting curve data for thalassemia diagnostics. Although we primarily trained and validated the models on large-scale datasets from a single clinical assay, the methodological framework itself is adaptable to other molecular diagnostic applications. This adaptability arises because the models operate directly on normalized melting curve features and Tm value encodings, independent of specific target sequences. Therefore, with appropriate retraining, this analytical framework can be deployed in applications such as infectious disease genotyping, mutation screening, methylation profiling [15], and high-resolution multiplex detection. The key requirement for such transfer lies in acquiring sufficient labeled melting curve data of the new assay to ensure that feature distributions are adequately represented during model training.

Our findings further underscore that the volume and balance of training data are major determinants of model performance, and accuracy plateaus once the models are exposed to sufficiently large datasets. When acquiring large-scale experimental data is infeasible, in silico simulation of melting curves and peak shapes can serve as an effective data augmentation strategy [16]. Such simulations can incorporate thermodynamic modeling of DNA duplex melting, stochastic noise injection, and baseline drift patterns, thereby mimicking the variability observed in real experiments. By generating synthetic but physically plausible melting curves, we can pre-train models on simulated data and then refine them on smaller experimental datasets. This approach could reduce the data requirements for applying ML-driven melting curve analysis in rare diseases or low-throughput research contexts.

A notable limitation of our current framework is its reduced performance when applied to unseen platforms. In our cross-platform validation [17], models trained on SLAN-96S data were tested using melting curves from a second instrument (Gentier 96R), and vice versa. Although peak temperatures remained largely consistent across systems, systematic differences in baseline noise and slope led to a marked decline in predictive accuracy. These observations emphasize the need for baseline harmonization or multi-platform data integration to ensure the model’s robustness across heterogeneous qPCR instruments.

Overall, our work establishes a robust foundation for scalable and platform-agnostic automated melting curve interpretation. By integrating large-scale data-driven learning, domain-specific in silico data augmentation, and cross-platform baseline harmonization techniques, we can extend the proposed framework beyond thalassemia molecular diagnostics to a broader range of Tm-based molecular assays. If further validated across diverse clinical contexts and qPCR platforms, this generalization has the potential to substantially reduce manual interpretation burden, improve inter-laboratory reproducibility, and accelerate the translation of high-throughput, ML-enhanced qPCR workflows into routine precision diagnostics.

## Data Availability

All data produced in the present study are available upon reasonable request to the authors.

## 5. Acknowledgments

This work was supported by the Sichuan Association for Promoting Science and Education under Grant No. KJXC24-0313.

